# *Are Our COVID Warriors Cared-for Enough?* A Nationwide Survey on Stress Among Doctors During the COVID-19 Pandemic

**DOI:** 10.1101/2020.07.03.20145748

**Authors:** Amritha Nair, Jagadeesh Menon, Ashwin Rammohan, Abdul R Hakeem, Sathya D Cherukuri, Naresh Shanmugam, Akila Rajakumar, Mettu S Reddy, Ilankumaran Kaliamoorthy, Mohamed Rela

## Abstract

Limited and uneven accessibility to healthcare is a major impediment in the fight against the COVID-19 pandemic which continues on inexorably, across various parts of the globe. We conducted a nationwide survey of a large sample of Indian doctors to measure levels of perceived stress, identify risk factors for severe stress and assess their response to current issues related to safety and well-being of the HCP community. The survey found severely stressed doctors to be younger (<45years), of female gender working in the ICU setting and insecure regarding their finances. Concern regarding PPE shortages and ethical dilemmas of rationing care are factors inducing severe stress amongst doctors working in ICU settings. This is the first such survey done in the context of the COVID-19 pandemic from the Indian sub-continent. The findings have important implications on the International healthcare community, especially across Africa, Asia & South America where the contagion continues to wreak havoc. The survey has identified factors which adversely impact the mental health of doctors during this Pandemic. This can act as a valuable guide for governmental authorities, professional organisations and hospital managements to establish support systems at multiple levels for these “COVID Warriors”.

## Introduction

The COVID-19 pandemic continues inexorably in India with infections crossing the half a million mark. Limited and uneven accessibility to healthcare in India is likely to be a major impediment in its fight against the pandemic ^1^. For health care professionals (HCP) at the forefront, safety of their families, increased working hours, expected salary cuts and projected PPE shortfalls are key issues. Public opinion in India towards doctors has always been ambivalent, and while there is widespread praise and support for their efforts, assaults on doctors and reports of doctors and families being ostracized for fear of spreading the infection have persisted. ^1–3^. Refusing to acknowledge and act on the warning signs of stress in this scenario can adversely affect a doctor’s professional, social and personal life. We conducted a nationwide survey of a large sample of Indian doctors to measure levels of perceived stress, identify risk factors for severe stress and assess their response to current issues related to safety and well-being of the HCP community.

## Methodology

An online survey using Google forms was conducted between 1^st^ May 2020 and 15^th^ May 2020. The survey questionnaire consisted of seven sections.(Supplement Table 1) The core component of the survey was the standardized Perceived Stress Scale (PSS-10) comprising 10 questions related to their feelings and thoughts during the last month answered on a Likert scale.^4^ Other sections of the survey included responder demographics, area of clinical work, extent of involvement in care of COVID-19 patients, possible causes of stress and techniques of destressing. The survey questionnaire was shared through email and Whatsapp and data was directly collected to an excel sheet for analysis.

## Results

520 doctors completed the questionnaire. 394 (76%) responders were under 45 years of age and 208 (40%) were female doctors. 90% of respondents had a post-graduate degree. 313 (60.2%) of the respondents were practicing in the private healthcare sector.

Based on the cumulative scores for the PSS-10 questionnaire, 109 (21%) had low stress, 371 (71%) had moderate stress and 40 (8%) reported severe perceived stress. On univariate analysis, factors associated with severe stress were female gender, age less than 45 years, junior doctors, doctors working in the ICU and doctors who reported being financially insecure. Multivariate analysis revealed female gender, being financially insecure and ICU as place of work as independent risk factors for severe stress. (Figure 1) Within the severe stress group, 45% of doctors reported ‘poor to terrible’ quality of sleep and 70% had considered seeking specialist counselling.

**Figure 1:**
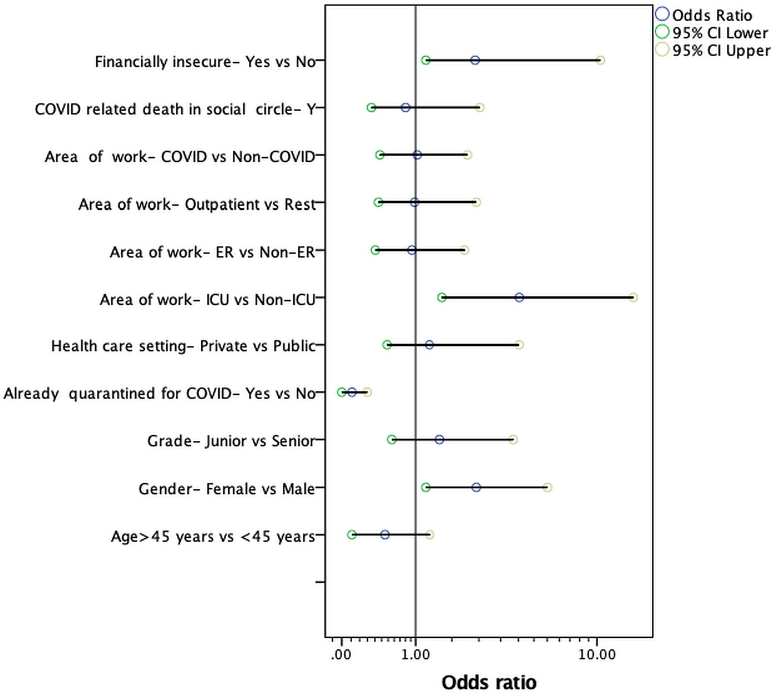
Forest Plot shows the odds ratios and 95% confidence intervals of factors predisposing to severe levels of perceived stress amongst 520 doctors responding to the survey. Logistic multivariate regression analysis with 1000 boot strapped samples was performed to identify significant factors and 95% confidence intervals. Female gender, ICU place of work and reported financial insecurity were significant risk factors (p<0.05).

## Discussion

Studies during the SARS & MERS outbreaks have reported higher stress among medical professionals including doctors^5,6^. The current pandemic is much larger in scale and has inflicted great damage even in countries with well developed healthcare systems. India with a mixed - predominantly privatised healthcare is likely to take a bigger hit.

Among the 520 doctors who responded to our survey, an alarming 4/5^th^ of the doctors were suffering from moderate or severe stress, of whom 70% perceived the need for counselling. Similar prevalence of stress among HCPs has been reported from other epicentres during this pandemic.^7–9^ Our survey has found severely stressed doctors to be younger (<45years), of female gender and insecure regarding their finances. In the Indian context, these would be doctors who are in the early phase of their careers with young families to support. They are the first to be affected financially by cut-backs in private hospitals secondary to substantial reduction in elective clinical work. Concern regarding PPE shortages and ethical dilemmas of rationing care in a pandemic situation are potential factors inducing severe stress amongst doctors working in ICU settings.^10^

This is the first such survey done in the context of the COVID-19 pandemic from the Indian sub-continent and has identified factors which have the potential to adversely impact the mental health of doctors. Despite the well-known limitations of such surveys, we believe that our findings are applicable to many countries in Asia and Africa with similar healthcare systems. Governmental authorities, professional organisations and hospital managements should take the lead and establish mechanisms at multiple levels to support doctors’ financial security, ensure adequate access to PPE at work and setup dedicated helplines and counselling facilities for those needing them. Ensuring their health is key to winning this war against COVID-19.

## Data Availability

On request via email from the corresponding author.

